# Structural covariance of early visual cortex is negatively associated with PTSD symptoms: A Mega-Analysis from the ENIGMA PTSD workgroup

**DOI:** 10.1101/2025.03.18.25324188

**Authors:** Nathaniel G. Harnett, Soumyaa Joshi, Poornima Kumar, Courtney Russell, Daniel G. Dillon, Justin T. Baker, Diego A. Pizzagalli, Milissa L. Kaufman, Lisa N. Nickerson, Neda Jahanshad, Lauren E. Salminen, Sophia I Thomopoulos, Jessie L. Frijling, Dick J. Veltman, Saskia B.J. Koch, Laura Nawijn, Mirjam van Zuiden, Ye Zhu, Gen Li, Jonathan Ipser, Xi Zhu, Orren Ravid, Sigal Zilcha-Mano, Amit Lazarov, Benjamin Suarez-Jimenez, Delin Sun, Ahmed Hussain, Ashley A. Huggins, Tanja Jovanovic, Sanne J.H. van Rooij, Negar Fani, Anna R. Hudson, Anika Sierk, Antje Manthey, Henrik Walter, Nic J.A. van der Wee, Steven J.A. van der Werff, Robert R.J.M. Vermeiren, Pavel Říha, Lauren A. M. Lebois, Isabelle M. Rosso, Elizabeth A. Olson, Israel Liberzon, Mike Angstadt, Seth G. Disner, Scott R. Sponheim, Sheri-Michelle Koopowitz, David Hofmann, Rongfeng Qi, Adi Maron-Katz, Austin Kunch, Hong Xie, Wissam El-Hage, Hannah Berg, Steven E. Bruce, Katie A. McLaughlin, Matthew Peverill, Kelly Sambrook, Marisa Ross, Ryan J. Herringa, Jack B. Nitschke, Richard J. Davidson, Terri A. deRoon-Cassini, Carissa W. Tomas, Jacklynn M. Fitzgerald, Jennifer Urbano Blackford, Bunmi O. Olatunji, Steven M. Nelson, Evan M. Gordon, Maria Densmore, Jean Théberge, Richard W.J. Neufeld, Miranda Olff, Li Wang, Dan J. Stein, Yuval Neria, Jennifer S. Stevens, Sven C. Mueller, Judith K. Daniels, Ivan Rektor, Anthony King, Nicholas D. Davenport, Thomas Straube, Guangming Lu, Amit Etkin, Xin Wang, Yann Quidé, Shmuel Lissek, Josh Cisler, Daniel W. Grupe, Christine Larson, Brandee Feola, Geoffrey May, Chadi G. Abdallah, Ruth Lanius, Paul M. Thompson, Rajendra A. Morey, Kerry Ressler

## Abstract

**Background:** Identifying robust neural signatures of posttraumatic stress disorder (PTSD) symptoms is important to facilitate precision psychiatry and help in understanding and treatment of the disorder. Emergent research suggests structural covariance of early visual regions is associated with later PTSD development. However, large-scale analyses are needed – in heterogeneous samples of trauma-exposed and trauma naive individuals – to determine if such a neural signature is a robust – and potentially a pretrauma – marker of vulnerability.

**Methods:** We analyzed data from the ENIGMA-PTSD dataset (n = 2,814) and the Human Connectome Project – Young Adult (HCP-YA) dataset (n = 890) to investigate whether structural covariance of early visual cortex is associated with either PTSD symptoms or perceived stress. Structural covariance was derived from a multimodal pattern previously identified in recent trauma survivors, and participant loadings on the profile were included in linear mixed effects models to evaluate associations with stress.

**Results:** Early visual cortex covariance loadings were negatively associated with PTSD symptoms in the ENIGMA-PTSD dataset. The relationship persisted when accounting for prior childhood maltreatment; supporting PTSD symptom specificity, no relationship was observed with depressive symptoms and no association was observed between loadings and perceived stress measures in the HCP-YA dataset.

**Conclusion:** Structural covariance of early visual cortex was robustly associated with PTSD symptoms across an international, heterogeneous sample of trauma survivors. Future studies should aim to identify specific mechanisms that underlie structural alterations in the visual cortex to better understand posttrauma psychopathology.

## Introduction

Posttraumatic stress disorder (PTSD) is a highly debilitating psychiatric condition. While trauma exposure is the principal antecedent of PTSD, not all trauma-exposed individuals go on to develop long-term posttraumatic dysfunction. Identification of robust signatures linked to increased risk for PTSD would help identify individuals most in need of resources to prevent the development of trauma-and stress-related psychopathology. Neuroimaging signatures may be valuable prognostic markers of future expression of symptoms in trauma-exposed individuals [1], however the robustness of previously observed associations may depend on the sample analyzed [2, 3]. Therefore, evaluation of potential neural signatures of PTSD in large, heterogeneous samples is needed for identification of generalizable neural markers of posttraumatic psychopathology, and determine if such brain metrics are viable targets for future treatment research.

Structures of the ventral visual stream represent one promising neural signature for predicting the onset of PTSD. Although a significant body of research implicates canonical threat neurocircuitry in PTSD psychopathology, growing evidence suggests that the ventral visual stream may be a key component in the development and maintenance of PTSD symptoms [4]. The ventral visual stream is a major visual processing pathway that plays a critical role in object recognition and perception [5, 6]. Prior reviews highlight that there are also reciprocal connections between canonical threat-related brain regions (e.g., amygdala and hippocampus) and regions of the ventral visual stream that help support emotional function [7]. PTSD has been associated with lower thickness of the occipital and inferior temporal cortex [8–10]. However, the association between visual circuit neurobiology and PTSD symptoms may vary with the timing of trauma exposure relative to neuroimaging. For example, prior research demonstrated that lower gray matter volume of the visual cortex was associated with PTSD symptoms and/or diagnosis months after trauma exposure [11]. In contrast, structural covariance of the ventral visual stream – particularly early visual regions such as V1 through V3 – was positively associated with PTSD symptom expression acutely after trauma exposure in two independent samples collected within approximately 2-4 weeks of trauma exposure [12, 13]. However, a greater delay (e.g., ∼6 months) between trauma exposure and brain imaging revealed a negative relationship between early visual structural covariance and PTSD symptoms [12]. Taken together, prior literature from our group and others suggests that PTSD is associated with alterations in the structure of early visual regions that may be partially dependent on the time since trauma exposure. Identifying whether the structure of early visual regions and the ventral visual stream are associated with PTSD in large, heterogeneous samples may yield generalizable neural signatures of posttraumatic dysfunction, and may help clarify how such a relationship unfolds over time.

The ENIGMA-PGC PTSD working group is an international consortium that collaboratively analyzes one of the largest neuroimaging datasets of trauma and PTSD to date. The pooling of data across countries and samples allows for “mega-analyses” of neuroimaging data for high-powered confirmatory and data-driven studies. Recent ENIGMA-PTSD mega-analyses found relationships between visual-system structures and PTSD symptoms using structural covariance networks (SCNs) generated from cortical morphometry [14, 15]. SCNs represent patterns of shared interregional variability within a participant that can be correlated with psychiatric phenotypes. Interestingly, lower covariance was observed in those with PTSD, compared to those without PTSD, in the visual system, including the anterior and middle occipital cortices. Furthermore, network properties (e.g., centrality) of regions along the ventral visual stream, such as the fusiform and occipital gyrus, varied as a function of PTSD status [16].

The prior findings suggest that structural covariance of early visual cortex may be a generalizable and robust marker for predicting PTSD symptoms. However, there have been limited investigations of out-of-sample, multimodal MRI-derived early visual SCNs, such as those identified previously [12, 13] in large samples of trauma/PTSD. SCNs derived from multimodal fusion methods can incorporate information across multiple MRI features in a manner that is not possible in univariate approaches [17]. Further, assessment of previously replicated SCNs in large and heterogeneous samples such as ENIGMA-PTSD would better elucidate the potential role of visual circuitry in the pathophysiology of PTSD. Of note, multimodal-derived early visual cortex SCNs and potential relationships with stress have not been investigated in trauma naive control samples. An association between a multimodal early visual SCN and general stress in typical or trauma naïve individuals may suggest that such an SCN is not PTSD-specific, and – if also confirmed in a PTSD sample – may be a potential pre-trauma marker of PTSD risk. Supporting a potential association in trauma naïve individuals, evidence suggests that visual pathway neurobiology is altered by experimental stress. Prior studies have found stress-related neural responses in visual regions, such as the lingual gyrus in humans [18], and stress-induced visual cortex atrophy in mice [19]. Identification of a marker of PTSD symptoms in trauma exposed, and heightened stress as a potential vulnerability marker in healthy individuals, may have significant translational benefit for the development of neuroscience-based predictive markers of psychopathology.

Therefore, the present study investigated associations between structural covariance networks of early visual cortex and posttraumatic stress in two large, independent datasets. We estimated individual participant loadings on a previously identified multimodal SCN overlapping the ventral visual stream in two datasets: 1) one consisting of previously trauma-exposed individuals and 2) a separate dataset of typical (healthy) young adults. We hypothesized that SCN loadings would be negatively associated with PTSD symptoms in the trauma sample – consistent with our prior investigations in later posttrauma phases. Further, we hypothesized that SCN loadings would be positively associated with a measure of perceived stress in neurotypical controls, in line with the potential for network covariance to be a pretrauma vulnerability factor.

## Methods and Materials

### Participants

Participants were drawn from two main data sources to define both trauma-exposed and trauma-naïve samples. Trauma-exposed participants’ data were provided by the ENIGMA-PGC PTSD working group. Broad aspects of imaging data sharing for the dataset have been previously described [10, 15, 16, 20, 21]. For the current analyses, data were available from 30 imaging sites across 9 countries from participants with and without PTSD (Table 1). Participants with available T1-weighted imaging data (see below) were included in the present analysis (n = 2,814). All study sites obtained local institutional review board or equivalent ethics committee approval, and all participants provided informed consent.

**Table 1.**
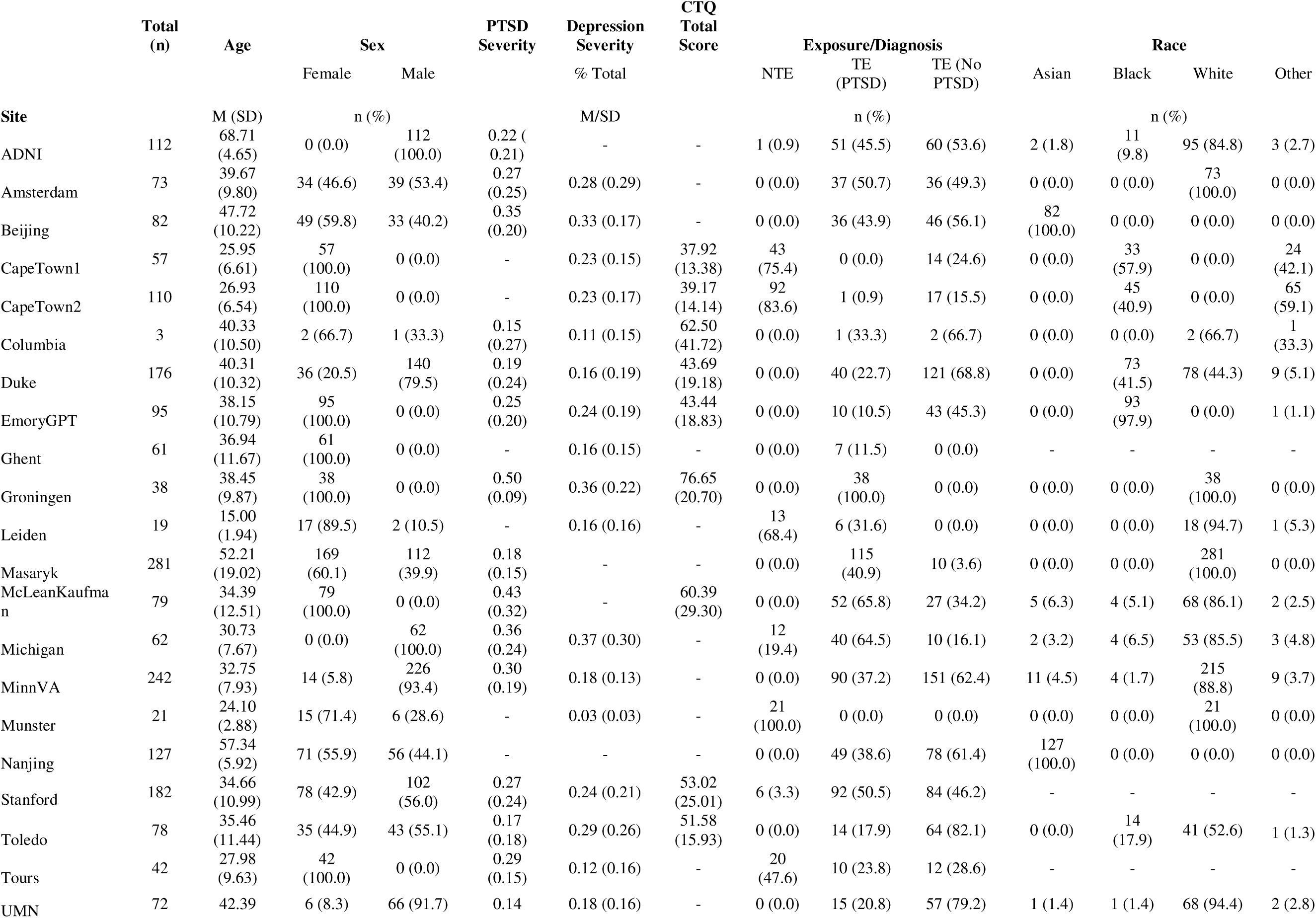
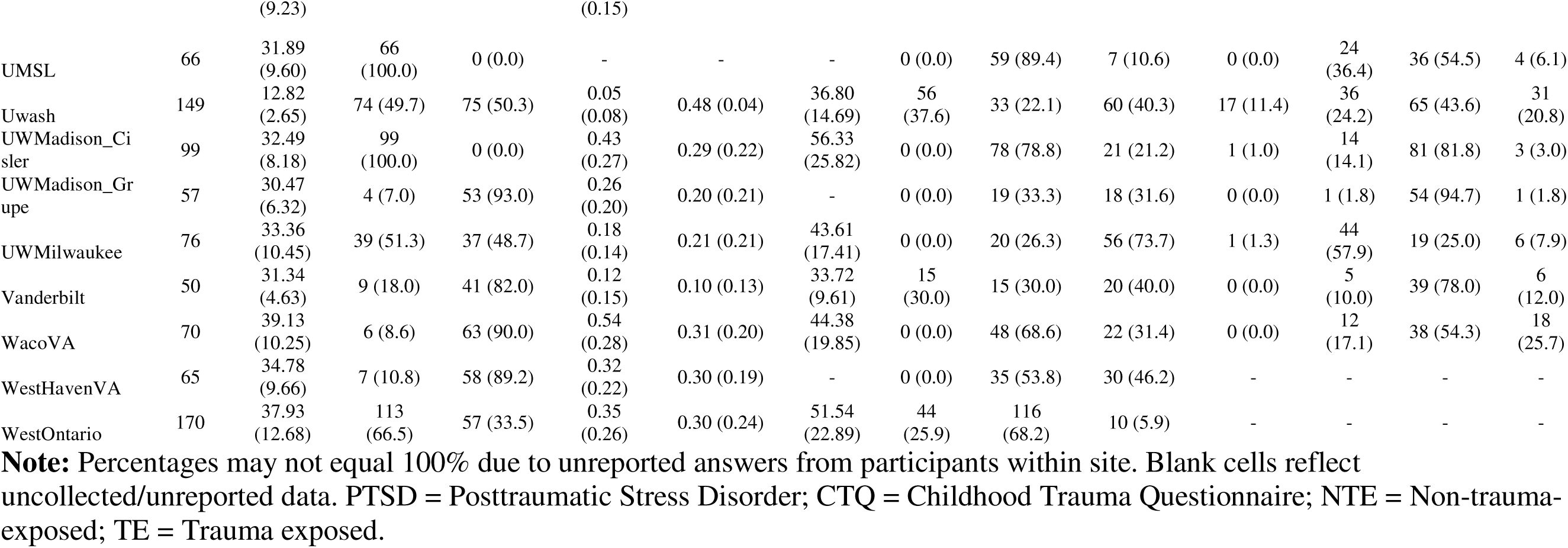
ENIGMA-PGC demographic data

An additional set of control participants was drawn from the Human Connectome Project Young Adult dataset (S1200 release). The HCP data were all collected at a single site on a single scanner that did not undergo any hardware/software upgrades during the data collection period. Details of participant recruitment have been previously documented [22]. For the present analyses, we selected data from individuals with T1-weighted and diffusion-weighted imaging from an ongoing multimodal data fusion analysis (n = 890). The HCP study was approved by the Washington University in St. Louis Human Research Protection Office (IRB #201204036, “Mapping the Human Connectome: Structure, Function, and Heritability”).

#### Demographics and Clinical Symptoms

In both datasets, participants reported their age and sex. Participants in the ENIGMA-PTSD dataset reported PTSD symptoms according to the DSM-IV or DSM-5 criteria depending on the study, through one of several different assessments (Table S1). A harmonized measure of symptom severity was obtained by calculating the percentage of total severity score endorsed by the participant relative to the possible total severity score of the assessment. Several sites also obtained measures of depression severity across several different questionnaires (Table S1), and a harmonized measure was determined by calculating the percentage of total severity relative to the possible total severity score, as above. Further, several sites collected data on childhood maltreatment using the Childhood Trauma Questionnaire (CTQ) [23] and the total CTQ score was included in our analyses. The ENIGMA-PTSD dataset contained trauma-exposed and trauma-naive participants. Given our hypothesis on the role of the ventral visual stream in understanding trauma-related responses, our primary analyses were restricted to trauma-exposed analyses in the ENIGMA-PTSD dataset. Follow-up analyses involving the trauma-naive group are noted in the results and described in detail in the supplement for completeness.

Participants in the HCP-YA dataset completed a battery of self-assessments related to mental health [17]. Given the dataset did not include typical trauma or PTSD measures (e.g., PCL-5), we selected more general “stress” measures for comparison, focusing on the “perceived stress” and Achenbach Adult Self-Report (ASR) scores. Perceived stress was operationalized as part of the NIH Toolbox Perceived Stress Survey using items from the Perceived Stress Scale [24]. Participants are asked about the degree to which events in the past month seemed unpredictable or uncontrollable and their coping resources on a Likert scale from 0-4. The total score was used as a measure of perceived stress. In follow-up analyses, we also considered scores from the Achenbach Adult Self-Report, administered as part of the NIH Toolbox [25]. As there was not a specific measure of PTSD symptoms included, we selected three ASR scale scores for comparison: Total scores, Anxiety/Depression scores, and Intrusive scores.

#### Magnetic resonance imaging

3D volumetric T1-weighted brain MRI data collected by participating sites was used to derive structural features for calculation of early visual structural covariance in both datasets, similar to our prior reports [12, 15, 16]. The previously observed SCN was predominantly composed of pial surface Area (PSA) and gray matter volume derived from voxel-based morphometry (VBM). For the ENIGMA-PTSD dataset, anatomical brain images were preprocessed at Duke University using the standardized ENIGMA 3.0 pipeline (https://enigma.ini.usc.edu/protocols/imaging-protocols/). Pial surface area across the cortex was extracted using FreeSurfer version 5.3 [26] and we applied a 10-mm Full-Width-at-Half-Max (FWHM) Gaussian kernel. Gray matter volume was calculated using FSLVBM [27] and data were smoothed using a 3-mm FWHM Gaussian kernel. The HCP-YA data were processed using the HCP minimal preprocessing pipeline, which has been previously described [28]. Cortical surface maps were reconstructed in FreeSurfer version 5.3 as part of the pipeline, and *fsaverage* resampled maps of PSA were smoothed with a 10-mm FWHM Gaussian kernel. As with the ENIGMA-PGC dataset, gray matter volume was calculated with FSLVBM and smoothed using a 3-mm FWHM Gaussian kernel.

Early visual structural covariance was calculated using previously applied approaches to back-project precomputed multimodal maps onto participant data [12, 29]. Specifically, 4D image files of participants’ VBM and PSA data (i.e., maps concatenated across participants) were created and used as inputs in dual regression. Previously identified early visual covariance maps from our prior report [12] were used as spatial maps to project onto participant data (Figure 1A). Component loadings from the first stage of dual regression were then averaged together and z-standardized to index individual participant early visual covariance loadings. The dual regression steps were separately applied for the ENIGMA-PTSD and HCP-YA datasets.

**Figure 1.**
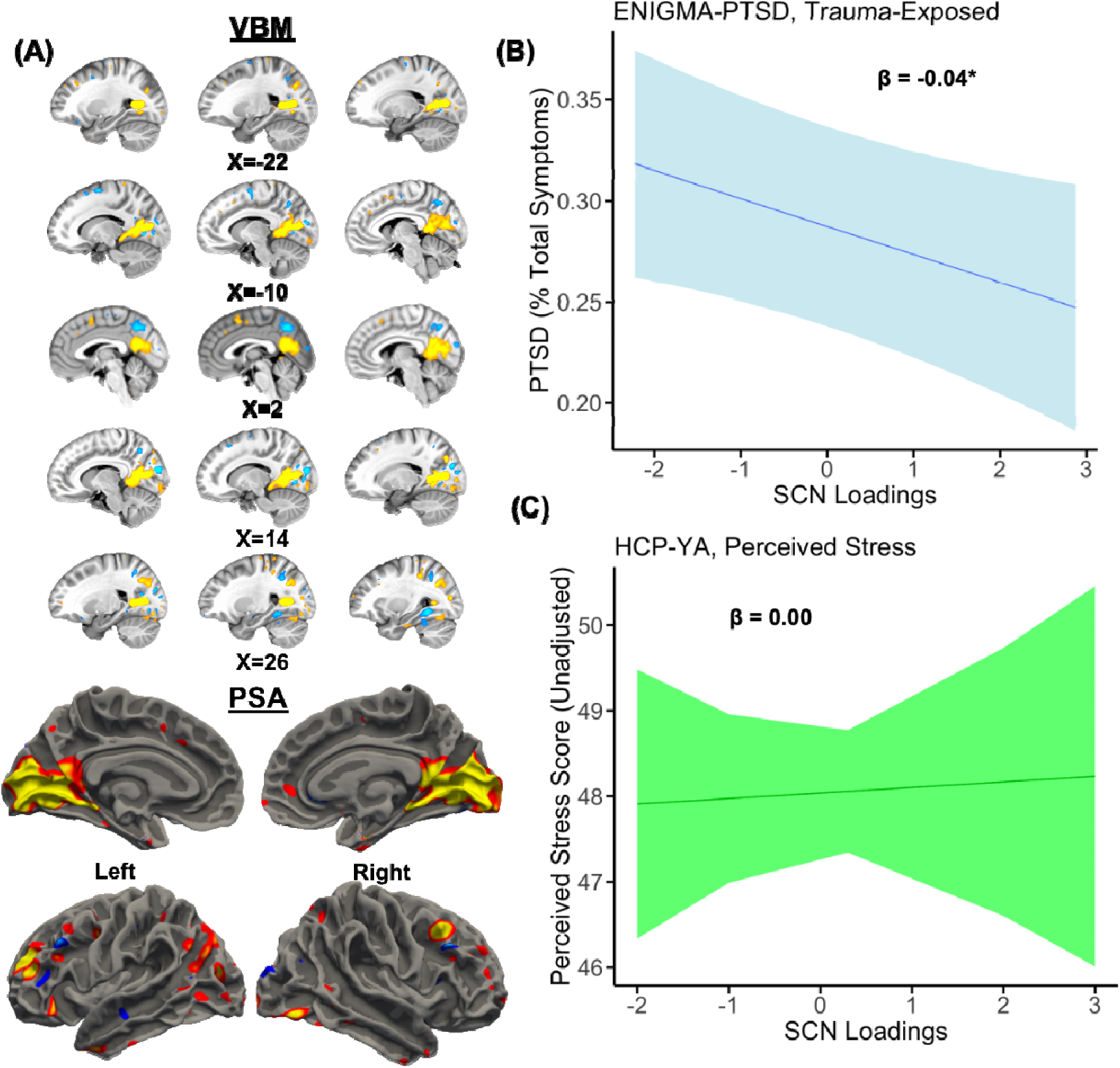
Early visual structural covariance is associated with PTSD symptoms in the ENIGMA-PGC cohort. Voxel-based morphometry (VBM) and pial surface area (PSA) features of a previously identified multimodal structural covariance network (SCN) [12] were projected onto data from the ENIGMA-PTSD and HCP-YA (A). Within the ENIGMA-PTSD dataset, individual participant loadings for the SCN were negatively correlated with PTSD symptom (B), however no relationship was observed between loadings and perceived stress in the HCP-YA dataset (C). Graphs represent the partial plots from linear mixed effects models. Solid lines represent the unique association from the model and the shaded bars represent the confidence intervals.

#### Statistical analyses

We completed linear mixed effect models to identify associations between early visual covariance loadings and PTSD symptoms. Participant age and sex were included as fixed-effect covariates in the model while participant site and scanner (ENIGMA-PTSD dataset) were included as random effects or Family ID (HCP-YA, to account for siblings/twins in the dataset); we also completed analyses with the HCP-YA dataset removing siblings in sensitivity analyses (supplemental material). Alternative techniques for harmonizing multisite neuroimaging data, such as ComBat-GAM [30], have previously been used with the present datasets [31]. However, ComBat-GAM typically requires high-dimensionality (i.e., multi-feature) data, whereas a benefit of the present approach is that multimodal covariance is reflected as a single variable (i.e., subject loading) for analysis.

Our primary models tested: 1) the association between SCN loadings and PTSD symptoms in trauma-exposed participants separately within the ENIGMA-PGC dataset, and 2) the association between SCN loadings and perceived stress within the HCP-YA dataset. Given our directional hypotheses (i.e., negative association between SCN loadings and PTSD symptoms for trauma-exposed, but positive for trauma-naive controls), for our primary analyses we employed one-tailed hypothesis testing with a nominal *p* < 0.05 as a significance threshold. The standardized beta for the linear mixed models were calculated using the *“effectsize”* package in R and 90% confidence intervals (CI) were calculated corresponding to our 1-tailed tests (specified in results).

We completed several follow-up and sensitivity analyses to understand the robustness of observed effects. For the ENIGMA-PGC dataset, we completed follow-up analyses to first investigate associations between SCN loadings and PTSD dimensions (i.e., percent severity for intrusion, avoidance, etc.). We completed additional analyses within the ENIGMA-PTSD dataset by removing participants with “zero” PTSD symptoms (i.e., symptoms assessed, but reported as zero) to control for potential zero-inflation of results. To test the specificity of our putative findings, we also investigated the association between SCN loadings and depression severity. Finally, additional sensitivity analyses determined if SCN loadings were associated with PTSD symptoms when covarying for CTQ scores in the subset of individuals with complete data.

## Results

### PGC-ENIGMA Dataset

Demographic information is presented in Table 1. We observed a negative relationship between SCN loadings and PTSD symptoms in trauma-exposed individuals with and without PTSD [β = -0.04, CI 90% = [-0.08, -0.01], p = 0.014 1-tailed, n = 2,027] (Figure 1B). Raw scatterplots between the independent and dependent variables are provided in the supplement (Figure S1A). Supplemental analyses of the trauma-naïve group revealed a positive association between SCN loadings and PTSD severity that was not significant when removing individuals with no PTSD symptoms (Supplemental Material).

Subscale analyses within the trauma-exposed group revealed SCN loadings were negatively associated with alterations in cognition and mood subscale scores (β = -0.06, CI 95% = [-0.11, -0.01], p = 0.029, n = 1,139). However, a significant relationship was not observed with intrusive (β = -0.02, CI 95% = [-0.07, 0.03], p = 0.422, n = 1,479), avoidance (β = -0.04, CI 95% = [-0.10, 0.02], p = 0.156, n = 1,140), or hyperarousal subscale scores (β = -0.04, CI 95% = [-0.09, 0.01], p = 0.110, n = 1,479).

The relationship between PTSD symptoms and SCN loadings remained significant (β = - 0.04, CI 90% = [-0.07, 0.00], p = 0.043, 1-tailed, n = 1,754) when removing individuals who reported no PTSD symptoms (n = 273) (Figure S1B). We also completed the same primary models described above using depression symptom severity and did not observe a significant association between SCN loadings and depression symptoms though the effect size was similar (β = -0.04, CI 95% = [-0.08, 0.01], p = 0.156, n = 1,386); an alternative model where depression severity was included as a covariate in models for PTSD severity revealed a similar effect to our main model [β = -0.03, CI 90% = [-0.06, 0.00], p = 0.038 1-tailed, n = 1,336]. An additional sensitivity analysis revealed SCN loadings remained associated with PTSD severity while covarying for CTQ total scores in a subset (n = 780) of participants (β = -0.07, CI 95% = [-0.13, -0.02], p = 0.005).

### HCP-YA Dataset

Demographic information on the sample is presented in Table 2. Our linear mixed effects models did not reveal a significant association between SCN loadings and perceived stress (β = 0.00, CI 90% = [-0.05, 0.06], p = 0.429, 1-tailed, n = 889) (Figure 1C). Follow-up analyses using the ASR did not show a relationship with ASR Total Scores (β = 0.05, CI 90% = [-0.01, 0.10], p = 0.092, 1-tailed, n = 887) (Figure S4). Subsequent analyses with ASR Anxiety/Depression (β = -0.01, CI 90% = [-0.07, 0.04], p = 0.369 1-tailed, n = 887) and ASR Intrusive scores (β = 0.03, CI 90% = [-0.03, 0.08], p = 0.208 1-tailed, n = 887) did not show significant relationships with SCN loadings. Removal of twins/siblings from the HCP-YA dataset did not impact the results (supplementary material). The findings suggest structural covariance of early visual regions is not associated with perceived stress in a typical control sample.

**Table 2.** HCP-YA demographic data

| Total (n) | 890 |
| --- | --- |
| *Age* | 28.71 (3.73) |
| ***Sex, n (%)*** |  |
| *Female* | 491 (55.2) |
| *Male* | 399 (44.8) |
| ***Race, n (%)*** |  |
| *Asian/Hawaiian/Pacific*<br>*Islander* | 57 ( 6.4) |
| *Black* | 122 (13.7) |
| *White* | 672 (75.5) |
| *Unknown* | 16 ( 1.8) |
| *Other* | 23 (2.6) |
| ***Perceived Stress, M (SD)*** | 48.03 (9.04) |
| ***Achenbach Self-report***<br>***(ASR), M (SD)*** |  |
| *Total* | 36.25 (22.18) |
| *Anxious/Depressed* | 5.75 (5.31) |
| *Intrusive* | 2.39 (2.15) |

## Discussion

Identifying robust and generalizable neuroimaging signatures of PTSD is essential for the development of neuroscience-informed predictive models and, ultimately, early interventions. Multimodal MRI-based indices of specific neural circuits, such as the ventral visual stream, may help in developing reproducible markers for PTSD symptoms. Here, we analyzed data from the ENIGMA-PGC PTSD Neuroimaging Working Group to investigate individual variability in a multimodal structural covariance network overlapping early visual brain regions that was related to PTSD symptom severity. Trauma-exposed individuals with and without PTSD showed a negative relationship between loadings on the network and PTSD symptoms, while trauma-naïve individuals showed a positive relationship (Supplemental Material). In a separate sample, the Human Connectome Project Young Adult cohort, there was no relationship between network loadings and more general measures of perceived stress. Taken together, our findings suggest that the decreased covariance of early visual regions may be associated with chronic PTSD symptom development.

The present findings partially replicate our prior reports of divergent associations between ventral visual stream network loadings and PTSD symptoms in acute versus chronic posttrauma samples [12, 13]. In our prior work, trauma survivors – with imaging data collected beyond the acute post-trauma period – showed a negative relationship between participant loadings on an SCN of early visual regions in the ventral visual stream and PTSD symptoms. Similarly, in the present heterogenous international sample, loadings of the same SCN were negatively associated with PTSD symptoms and in particular negative alterations in cognition and mood. A growing body of research suggests that trauma exposure is related to altered function and structure along the ventral visual stream which is, in turn, related to posttraumatic dysfunction [9, 15, 32]. Further, the present finding of chronic PTSD symptom severity associated with early visual cortex covariance mirrors our prior work from the AURORA study [12]. The current results suggest that weakened covariance of the ventral visual stream, in the chronic post-trauma phase, may partially underlie the development of affective trauma-related cognitions potentially through disruption of connections with canonical emotion regulation regions though additional research is needed.

A speculative mechanism underlying visual pathway alterations in PTSD is that the stress of trauma exposure or PTSD itself induces excitotoxic mechanisms that contribute to overall weakening of stimulus processing pathways. For example, traumatic stress or PTSD-related stressors may increase excitatory neurotransmitter release, and/or potentiate activity in circuits important for emotional processing (e.g., threat/visual regions). Previous reviews highlight that–across both preclinical and human studies– traumatic stress may lead to increased glutamate and decreased GABA in threat and sensorial regions [33]. For example, previous work in trauma survivors observed greater glutamate/glutamine within the prefrontal cortex was associated greater acute PTSD symptoms [34]. Disrupted glutamatergic/GABAergic processes, or sustained activation of visual/threat circuitry (e.g., heightened attention processes) may, in turn, contribute to degradation of the underlying structural connections between regions. In line with such reasoning, we found that lower SCN loadings were associated with greater PTSD symptoms, which may reflect degradation of structural integrity of the ventral visual stream across VBM and PSA features. Further, while childhood maltreatment and exposure to traumatic events are known to be related to diminished integrity and volume of visual pathways [35–37], our findings persisted when covarying for childhood maltreatment. Thus, while prior childhood trauma may contribute to changes in visual circuit biology, variability in early visual cortex covariance may be specific to PTSD symptoms rather than trauma itself though prior work suggests timing may play a role as well. Taken together, our findings suggest that the structure of the ventral visual stream plays a potentially important role in the maintenance and severity of PTSD symptoms after trauma exposure.

Contrary to our hypothesis, we did not observe an association between SCN loadings and perceived stress among participants within the HCP-YA dataset. Previous research in healthy participants suggests that visual cortex and the ventral visual stream are involved in responses to stressors, although the majority of such work assessed brain function. For example, recent work demonstrated that neuromodulation of the visual cortex reduced the intensity of experimentally-induced intrusive memories in healthy controls [38]. Further, previous work demonstrates that levels of social fearfulness in healthy participants is associated with slower habituation of neural responses to facial stimuli within visual cortex [39]. The findings may suggest that function of the visual cortex may play a more significant role in response to stress. However, there are limited data on brain structural associations with perceived stress in healthy samples. Evidence from animal models suggests that severe stress can contribute to atrophy within the visual cortex and related regions [19], and recent work in humans suggests stress can induce rapid changes in gray matter volume [40]. However, there is limited comparative or longitudinal work in trauma-exposed individuals showing that traumatic stress may acutely alter visual cortex structure. It is thus unclear from the present findings if the current SCN is an actionable pre-trauma marker of PTSD susceptibility. The lack of association in the HCP-YA dataset may suggest exposure to traumatic events is a necessary condition for the predictive utility of the network. Alternatively, it may suggest that the association is specific to PTSD symptoms and that measures of perceived stress do not capture the same process that underlies visual stream associations with trauma exposure. Our supplementary analyses of the NTE group partially support such an interpretation, since we observed a positive relationship between PTSD symptoms and SCN loadings among NTE individuals in the ENIGMA-PGC dataset. However, very few NTE participants reported experiencing PTSD symptoms (n = 12) and no significant association was observed in the reduced sample. There is also a lack of clarity around what endorsement of PTSD symptoms means when individuals do not have a history of trauma exposure. The present results suggest that additional work is needed to determine if the SCN is a useful marker for pre-trauma identification. One potential approach is to leverage naturalistic longitudinal datasets, such as the UK Biobank [38], to select a subset of individuals who experience trauma during the study period and determine if the SCN is then associated with later PTSD symptoms.

There are several limitations to consider when interpreting the present results. While the ENIGMA-PGC is the largest consortium sample of trauma-related MRI to date, there is significant heterogeneity in the assessment of PTSD symptoms, collection of data, inclusion of pretraumatic factors, and demographic data availability that may moderate the present findings. Further, there is evidence to suggest that additional stressors experienced before the index trauma, such as childhood maltreatment, have observable associations with sensorial neurobiology in adult trauma survivors [36, 41]. Although the early visual SCN was associated with PTSD symptoms while covarying for childhood maltreatment, there were a limited number of individuals with complete CTQ data and further consideration of potential associations with pretraumatic stressors is needed. It should also be noted that, although we had directional *a priori* hypotheses based on prior work, the observed effect sizes in the present analyses are small. Small effects may reflect some of the heterogeneity in the present sample or suggest the relative influence of visual circuit covariance on PTSD is weak. However, the replication of our prior work – including subscale specificity in the association – suggests early visual cortex covariance may still be a promising neural signature for PTSD. Further, such small effects may still be clinically meaningful if a robust neural signature can be translated to effective therapeutics. Another consideration is the relatively limited availability of data from other modalities (e.g., diffusion imaging) to reconstruct the SCN for each participant. Analytic limitations in consortium studies can preclude inclusion of MRI features and contribute to reduced statistical power. Further research including other modalities in similarly large samples may be helpful for better understanding ventral visual stream covariance in PTSD.

In conclusion, the present analyses revealed that structural covariance of early visual brain regions is negatively associated with PTSD symptoms in an international sample of trauma-exposed individuals. Critically, the structural covariance of the network is not associated with individual variability in general stress in a typical control sample, suggesting it is not a pre-trauma marker of PTSD development. Trauma exposure may therefore be a prerequisite for structural disruption of the visual pathway that contributes to PTSD, which may be due to the unique stress of trauma. The findings highlight an important role for the structure of visual processing pathways in understanding the neuroetiology of trauma and stress-related disorders.

## Data Availability

The Human Connectome Project data are available as part of approved access to the NIMH Data Archive. The ENIGMA-PTSD data are available upon request to the ENIGMA-PGC PTSD Working Group.

## Acknowledgements

The present work was funded by: National Institute of Mental Health (K01MH129828) and the Brain Behavior Research Foundation (NGH); R21MH112956, R01MH119227, McLean Hospital Trauma Scholars Fund, Barlow Family Fund, Julia Kasaparian Fund for Neuroscience Research; R01MH111671, R01MH117601, R01AG059874, MJFF 14848; ZonMw, the Netherlands organization for Health Research and Development (40-00812-98-10041), and the Academic Medical Center Research Council (110614); the National Natural Science Foundation of China (No. U21A20364 and No. 31971020), the Key Project of the National Social Science Foundation of China (No. 20ZDA079), the Key Project of Research Base of Humanities and Social Sciences of Ministry of Education (No.16JJD190006), and the Scientific Foundation of Institute of Psychology, Chinese Academy of Sciences (No. E2CX4115CX); NARSAD 27040; NIMH K01 MH122774; NIMH K01 MH118428-01; RO1 MH111671; VISN6 MIRECC; MH098212; MH071537; M01RR00039; UL1TR000454; HD071982; HD085850; Narsad Young Investigator; K01MH121653; MH101380; German Research Foundation grant to J. K. Daniels (numbers DA 1222/4-1 and WA 1539/8-2); the Ministry of Health of the Czech Republic, grant no. AZV NV18-7 04-00559, MAFIL supported by the Czech-BioImaging large RI project (LM2023050 funded by MEYS CR); K23MH112873; R01MH113574; VA RR&D 1K1RX002325; 1K2RX002922; VA RR&D I01RX000622; CDMRP W81XWH-08–2–0038; German Research Society (Deutsche Forschungsgemeinschaft, DFG; SFB/TRR 58: C06, C07); The Natural Science Foundation of Jiangsu Province (No. BK20221554), and the Foundation for the Social Development Project of Jiangsu (No. BE2022705); PHRC, Fondation Pierre Deniker and SFR FED4226; R01-MH132221, R01-MH128371; Dana Foundation (to Dr. Nitschke); the University of Wisconsin Institute for Clinical and Translational Research (to Dr. Emma Seppala); a National Science Foundation Graduate Research Fellowship (to Dr. Grupe); the National Institute of Mental Health (NIMH) R01-MH043454 and T32-MH018931 (to Dr. Davidson); and a core grant to the Waisman Center from the National Institute of Child Health and Human Development (P30-HD003352); R01 MH106574; R21MH106998; VA CSR&D 1IK2CX001680; VISN17 Center of Excellence Pilot funding; the SAMRC Unit on Risk & Resilience in Mental Disorders; R01MH105355-01A; grant 01J05415 from the Special Research Fund (BOF) at Ghent University; VA RR&D 1IK2RX000709; 1R01MH110483 and 1R21MH098198; VA National Center for PTSD; The Beth K and Stuart Yudofsky Chair in the Neuropsychiatry of Military Post Traumatic Stress Syndrome; Department of Defense award number W81XWH-12-2-0012; ENIGMA was also supported in part by NIH U54 EB020403 from the Big Data to Knowledge (BD2K) program, R56AG058854, R01MH116147, R01MH111671, and P41 EB015922; RO1 MH111671; VISN6 MIRECC

## References

1. Hinojosa CA, George GC, Ben-Zion Z. Neuroimaging of posttraumatic stress disorder in adults and youth: progress over the last decade on three leading questions of the field. Molecular Psychiatry 2024 29:10. 2024;29:3223–3244.

2. Ben-Zion Z, Spiller TR, Keynan JN, Admon R, Levy I, Liberzon I, et al. Evaluating the Evidence for Brain-Based Biotypes of Psychiatric Vulnerability in the Acute Aftermath of Trauma. American Journal of Psychiatry. 2023;180:146–154.

3. Stevens JS, Harnett NG, Lebois LAM, Van Rooij JH, Ely TD, Roeckner A, et al. Brain-Based Biotypes of Psychiatric Vulnerability in the Acute Aftermath of Trauma. Am J Psychiatry. 2021;178:1037–1049.

4. Harnett NG, Fleming LL, Clancy KJ, Ressler KJ, Rosso IM. Affective visual circuit dysfunction in trauma and stress-related disorders. Biol Psychiatry. 2024. 10 July 2024. 10.1016/J.BIOPSYCH.2024.07.003.

5. Long B, Yu CP, Konkle T. Mid-level visual features underlie the high-level categorical organization of the ventral stream. Proc Natl Acad Sci U S A. 2018;115:E9015–E9024.

6. Kravitz DJ, Saleem KS, Baker CI, Ungerleider LG, Mishkin M. The ventral visual pathway: An expanded neural framework for the processing of object quality. Trends Cogn Sci. 2013;17:26–49.

7. Pessoa L, Adolphs R. Emotion processing and the amygdala: From a ‘low road’ to ‘many roads’ of evaluating biological significance. Nat Rev Neurosci. 2010;11:773–782.

8. Wrocklage KM, Averill LA, Scott JC, Averill CL, Schweinsburg B, Trejo M, et al. Cortical thickness reduction in combat exposed U . S . veterans with and without PTSD. European Neuropsychopharmacology. 2017;27:515–525.

9. Crombie KM, Ross MC, Letkiewicz AM, Sartin-Tarm A, Cisler JM. Differential relationships of PTSD symptom clusters with cortical thickness and grey matter volumes among women with PTSD. Sci Rep. 2021;11:1825.

10. Wang X, Xie H, Chen T, Cotton AS, Salminen LE, Logue MW, et al. Cortical volume abnormalities in posttraumatic stress disorder: an ENIGMA-psychiatric genomics consortium PTSD workgroup mega-analysis. Mol Psychiatry. 2021;26:4331–4343.

11. Cwik JC, Vahle N, Woud ML, Potthoff D, Kessler H, Sartory G, et al. Reduced gray matter volume in the left prefrontal, occipital, and temporal regions as predictors for posttraumatic stress disorder: a voxel-based morphometric study. Eur Arch Psychiatry Clin Neurosci. 2019. 2019. 10.1007/s00406-019-01011-2.

12. Harnett NG, Finegold KE, Lebois LAM, van Rooij SJH, Ely TD, Murty VP, et al. Structural covariance of the ventral visual stream predicts posttraumatic intrusion and nightmare symptoms: a multivariate data fusion analysis. Transl Psychiatry. 2022;12:321.

13. Harnett NG, Stevens JS, Fani N, van Rooij SJH, Ely TD, Michopoulos V, et al. Acute Posttraumatic Symptoms Are Associated With Multimodal Neuroimaging Structural Covariance Patterns: A Possible Role for the Neural Substrates of Visual Processing in Posttraumatic Stress Disorder. Biol Psychiatry Cogn Neurosci Neuroimaging. 2020. 2020. 10.1016/j.bpsc.2020.07.019.

14. Sun D, Haswell CC, Morey RA, De Bellis MD. Brain structural covariance network centrality in maltreated youth with PTSD and in maltreated youth resilient to PTSD. Dev Psychopathol. 2019;31:557–571.

15. Sun D, Rakesh G, Clarke-Rubright EK, Haswell CC, Logue MW, O’Leary EN, et al. Remodeling of the Cortical Structural Connectome in Posttraumatic Stress Disorder: Results From the ENIGMA-PGC Posttraumatic Stress Disorder Consortium. Biol Psychiatry Cogn Neurosci Neuroimaging. 2022;7:935–948.

16. Rakesh G, Logue MW, Clarke-Rubright E, Haswell CC, Thompson PM, De Bellis MD, et al. Network Centrality and Modularity of Structural Covariance Networks in Posttraumatic Stress Disorder: A Multisite ENIGMA-PGC Study. Brain Connect. 2023;13:211–225.

17. Calhoun VD, Sui J. Multimodal Fusion of Brain Imaging Data: A Key to Finding the Missing Link(s) in Complex Mental Illness. Biol Psychiatry Cogn Neurosci Neuroimaging. 2016;1:230–244.

18. Wheelock MD, Harnett NG, Wood KH, Orem TR, Granger DA, Mrug S, et al. Prefrontal cortex activity is associated with biobehavioral components of the stress response. Front Hum Neurosci. 2016;10:222304.

19. Yoshii T, Oishi N, Ikoma K, Nishimura I, Sakai Y, Matsuda K, et al. Brain atrophy in the visual cortex and thalamus induced by severe stress in animal model. Sci Rep. 2017;7:12731.

20. Logue MW, Amstadter AB, Baker DG, Duncan L, Koenen KC, Liberzon I, et al. The Psychiatric Genomics Consortium Posttraumatic Stress Disorder Workgroup: Posttraumatic Stress Disorder Enters the Age of Large-Scale Genomic Collaboration. Neuropsychopharmacology. 2015;40:2287–2297.

21. Dennis EL, Disner SG, Fani N, Salminen LE, Logue M, Clarke EK, et al. Altered white matter microstructural organization in posttraumatic stress disorder across 3047 adults: results from the PGC-ENIGMA PTSD consortium. Mol Psychiatry. 2021;26:4315–4330.

22. Van Essen DC, Ugurbil K, Auerbach E, Barch D, Behrens TEJ, Bucholz R, et al. The Human Connectome Project: A data acquisition perspective. Neuroimage. 2012;62:2222.

23. Bernstein DP, Stein JA, Newcomb MD, Walker E, Pogge D, Ahluvalia T, et al. Development and validation of a brief screening version of the Childhood Trauma Questionnaire. Child Abuse Negl. 2003;27:169–190.

24. Kupst MJ, Butt Z, Stoney CM, Griffith JW, Salsman JM, Folkman S, et al. Assessing Stress and Self-Efficacy for the NIH Toolbox for Neurological and Behavioral Function. Anxiety Stress Coping. 2015;28:531.

25. Hodes RJ, Insel TR, Landis SC, NIH Blueprint for Neuroscience Research. The NIH Toolbox: Setting a standard for biomedical research. Neurology. 2013;80:S1.

26. Dale AM, Fischl B, Sereno MI. Cortical surface-based analysis: I. Segmentation and surface reconstruction. Neuroimage. 1999;9:179–194.

27. Smith SM, Jenkinson M, Woolrich MW, Beckmann CF, Behrens TEJ, Johansen-Berg H, et al. Advances in functional and structural MR image analysis and implementation as FSL. Neuroimage, vol. 23, 2004.

28. Glasser MF, Sotiropoulos SN, Wilson JA, Coalson TS, Fischl B, Andersson JL, et al. The minimal preprocessing pipelines for the Human Connectome Project. Neuroimage. 2013;80:105–124.

29. Li H, Smith SM, Gruber S, Lukas SE, Silveri MM, Hill KP, et al. Denoising scanner effects from multimodal MRI data using linked independent component analysis. Neuroimage. 2020;208.

30. Pomponio R, Erus G, Habes M, Doshi J, Srinivasan D, Mamourian E, et al. Harmonization of large MRI datasets for the analysis of brain imaging patterns throughout the lifespan. Neuroimage. 2020;208:116450.

31. Sun D, Rakesh G, Haswell CC, Logue M, Baird CL, O’Leary EN, et al. A comparison of methods to harmonize cortical thickness measurements across scanners and sites. Neuroimage. 2022;261.

32. Harnett NG, van Rooij SJH, Ely TD, Lebois LAM, Murty VP, Jovanovic T, et al. Prognostic neuroimaging biomarkers of trauma-related psychopathology: resting-state fMRI shortly after trauma predicts future PTSD and depression symptoms in the AURORA study. Neuropsychopharmacology. 2021;46:1263–1271.

33. Averill LA, Purohit P, Averill CL, Boesl MA, Krystal JH, Abdallah CG. Glutamate Dysregulation and Glutamatergic Therapeutics for PTSD: Evidence from Human Studies. Neurosci Lett. 2017;649:147.

34. Harnett NG, Wood KH, Ference EW, Reid MA, Lahti AC, Knight AJ, et al. Glutamate/glutamine concentrations in the dorsal anterior cingulate vary with Post-Traumatic Stress Disorder symptoms. J Psychiatr Res. 2017;91:169–176.

35. Tomoda A, Polcari A, Anderson CM, Teicher MH. Reduced Visual Cortex Gray Matter Volume and Thickness in Young Adults Who Witnessed Domestic Violence during Childhood. PLoS One. 2012;7:e52528.

36. Teicher MH, Samson JA, Anderson CM, Ohashi K. The effects of childhood maltreatment on brain structure, function and connectivity. Nature Reviews Neuroscience 2016 17:10. 2016;17:652–666.

37. Choi J, Jeong B, Polcari A, Rohan ML, Teicher MH. Reduced fractional anisotropy in the visual limbic pathway of young adults witnessing domestic violence in childhood. Neuroimage. 2012;59:1071–1079.

38. Herz N, Bar-Haim Y, Tavor I, Tik N, Sharon H, Holmes EA, et al. Neuromodulation of Visual Cortex Reduces the Intensity of Intrusive Memories. Cerebral Cortex (New York, NY). 2022;32:408.

39. Avery SN, Blackford JU. Slow to warm up: the role of habituation in social fear. Soc Cogn Affect Neurosci. 2016;11:1832.

40. Uhlig M, Reinelt JD, Lauckner ME, Kumral D, Schaare HL, Mildner T, et al. Rapid volumetric brain changes after acute psychosocial stress. Neuroimage. 2023;265:119760.

41. Wong SA, Lebois LAM, Ely TD, van Rooij SJH, Bruce SE, Murty VP, et al. Internal capsule microstructure mediates the relationship between childhood maltreatment and PTSD following adulthood trauma exposure. Molecular Psychiatry 2023. 2023:1–10.

