## Supplemental Material for "Structural covariance of early visual cortex is negatively associated with PTSD symptoms: A Mega-Analysis from the ENIGMA PTSD workgroup"

Harnett et al.

**Supplementary Results**

*ENIGMA-PGC: NTE Participants*

Supplementary analyses with the NTE group revealed a positive association between PTSD symptoms and SCN loadings in the NTE group (β = 0.12, CI 90% = [0.01, 0.24], p = 0.039 1-tailed) (Figure S2A-B). However, a large proportion of NTE participants had no PTSD symptoms (i.e., 0s) which may contribute to zero inflation (n = 122) as only 12 participants had PTSD symptoms. Repeating the analyses when excluding individuals with no reported PTSD symptoms, we did not observe a significant relationship between SCN loadings and PTSD symptoms (β = 0.39, CI 90% = [-0.29, 1.06], p = 0.142, 1-tailed) (Figure S2C). SCN loadings did not differ between NTE and TE participants either with (β = 0.02, CI 95% = [-0.16, 0.19], p = 0.850) or without (β = 0.10, CI 95% = [-0.08, 0.28], p = 0.285) PTSD (Figure S3).

*HCP-YA Dataset (Non-twins)*

We performed supplementary analyses with monozygotic and dizygotic twins excluded (n = 435) equivalent to our primary analyses. Linear mixed effects models did not reveal a significant association between SCN loadings and perceived stress (β = 0.00, CI 90% = [-0.07, 0.09], p = 0.429, 1-tailed, n = 434). Similarly, there was no relationship between the SCN and ASR Total Scores (β = 0.06, CI 90% = [-0.02, 0.13], p = 0.123, 1-tailed, n = 434), Anxiety/Depression (β = 0.00, CI 90% = [-0.08, 0.08], p = 0.499, 1-tailed, n = 434), or Intrusive scores (β = 0.00, CI 90% = [-0.07, 0.09], p = 0.440, 1-tailed, n = 434).

**Table S1. PTSD and Depression measures across sites**

| Site | PTSD Severity | PTSD Diagnosis | Depression |
| --- | --- | --- | --- |
| ADNI | CAPS4 | CAPS4 | GDS_SF |
| Amsterdam | CAPS4 | CAPS4 | HADS_D |
| Beijing | PCL5 | PCL5 | CES_D |
| CapeTown1 | NA | MINI | BDI |
| CapeTown2 | NA | MINI | BDI |
| Columbia | CAPS5 | CAPS5 | HAM_D |
| Duke | CAPS4, CAPS5, DTS | CAPS4, CAPS5, DTS | BDI |
| EmoryGTP | CAPS4, MPSS | CAPS4 | BDI |
| Ghent | NA | MINI | BDI |
| Groningen | CAPS4 | CAPS4 | BDI |
| Leiden | NA | ADIS-C/P | CDI |
| Masaryk | PCL-C | PCL-C | GDS_SF |
| McLeanKaufman | CAPS5 | CAPS5 | NA |
| Michigan | CAPS4 | CAPS4 | DASS21 |
| MinnVA | CAPS4 | CAPS4 | BDI |
| Munster | NA | SCID4 | BDI |
| Nanjing | CAPS4 | SCID4 | SCID4 |
| Stanford | CAPS4 | CAPS4 | BDI |
| Toledo | CAPS4 | CAPS4 | CES_D, DASS21 |
| Tours | CAPS4 | CAPS4 | BDI_SF |
| UMN | CAPS4 | CAPS4 | BDI |
| UMSL | NA | CAPS4 | NA |
| UWash | CAPS5 | CAPS4, CAPS5 | CDI_2 |
| UWMadison_Cisler | PCLC, PCL5 | CAPS5, SCID4 | BDI |
| UWMadison_Grupe | CAPS4 | CAPS4 | BDI |
| UWMilwaukee | CAPS5 | CAPS5 | DASS21 |
| Vanderbilt | CAPS5 | CAPS5 | BDI |
| WacoVA | PCL5 | PCL5 | BDI |
| WestHavenVA | CAPS4 | CAPS4 | BDI |
| WestOntario | CAPS4, CAPS5, DTS | CAPS4, CAPS5 | DBI |

**Figure S1.** Scatterplots of early visual structural covariance network (SCN) loadings and PTSD symptoms in the ENIGMA-PTSD dataset including (A) and excluding (B) individuals with zero PTSD symptoms, as well as associations with perceived stress in the HCP-YA dataset (C). Graphs represent the scatter plots for the independent variables. Solid lines represent the linear line of best fit, the shaded bars represent the confidence intervals, and blue dots represent the individual data points.

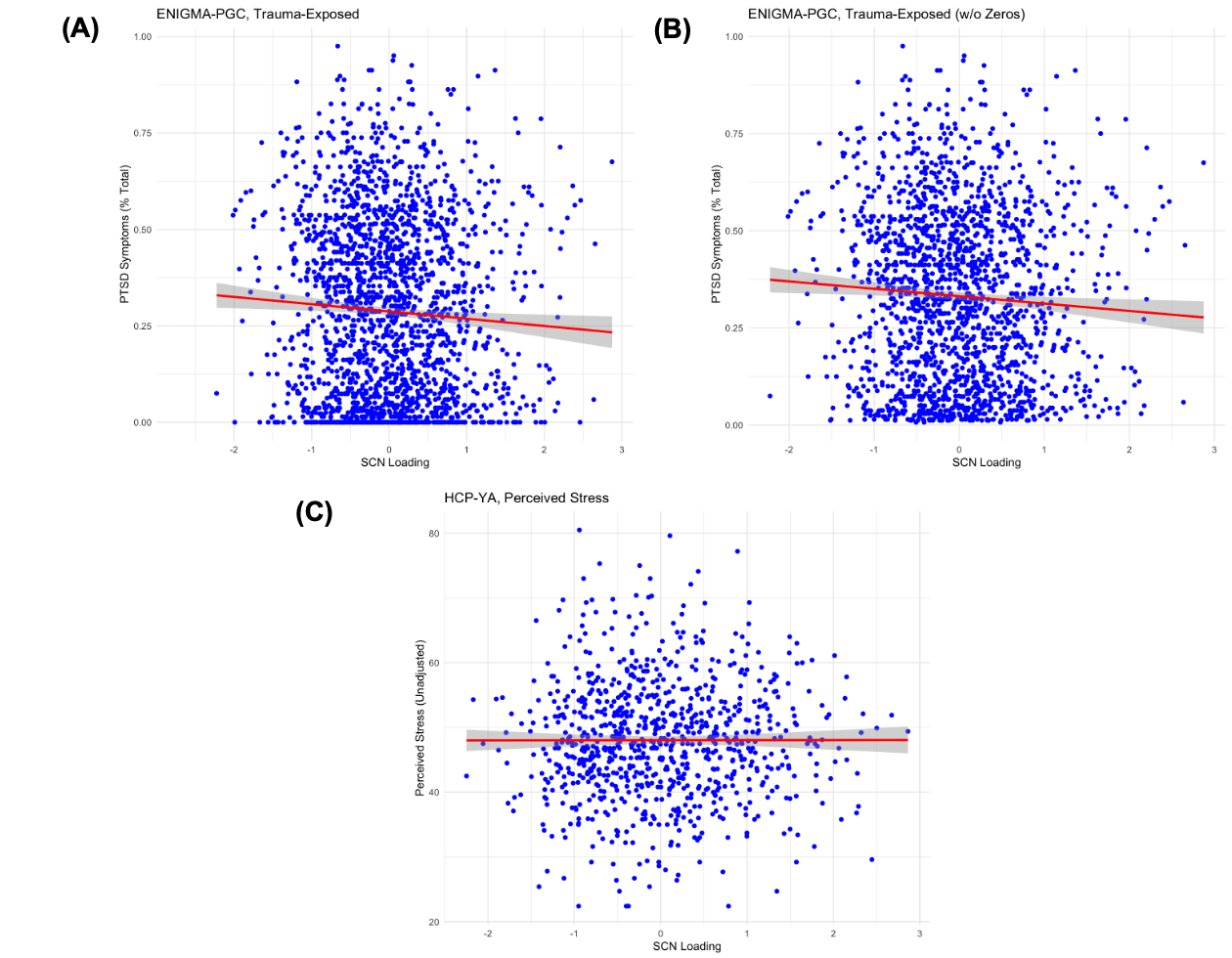

**Figure S2.** Early visual structural covariance network (SCN) loadings and PTSD symptoms in non-trauma-exposed (NTE) participants in the ENIGMA-PTSD dataset. The graph in (A) represents the partial plots from linear mixed effects models. Solid lines represent the unique association from the model and the shaded bars represent the confidence intervals. The graph in (B) represents the scatter plot for the independent variables. Solid lines represent the linear line of best fit, the shaded bars represent the confidence intervals, and blue dots represent the individual data points. The graph in (C) represents the same scatter plot removing individuals with zero PTSD symptoms.

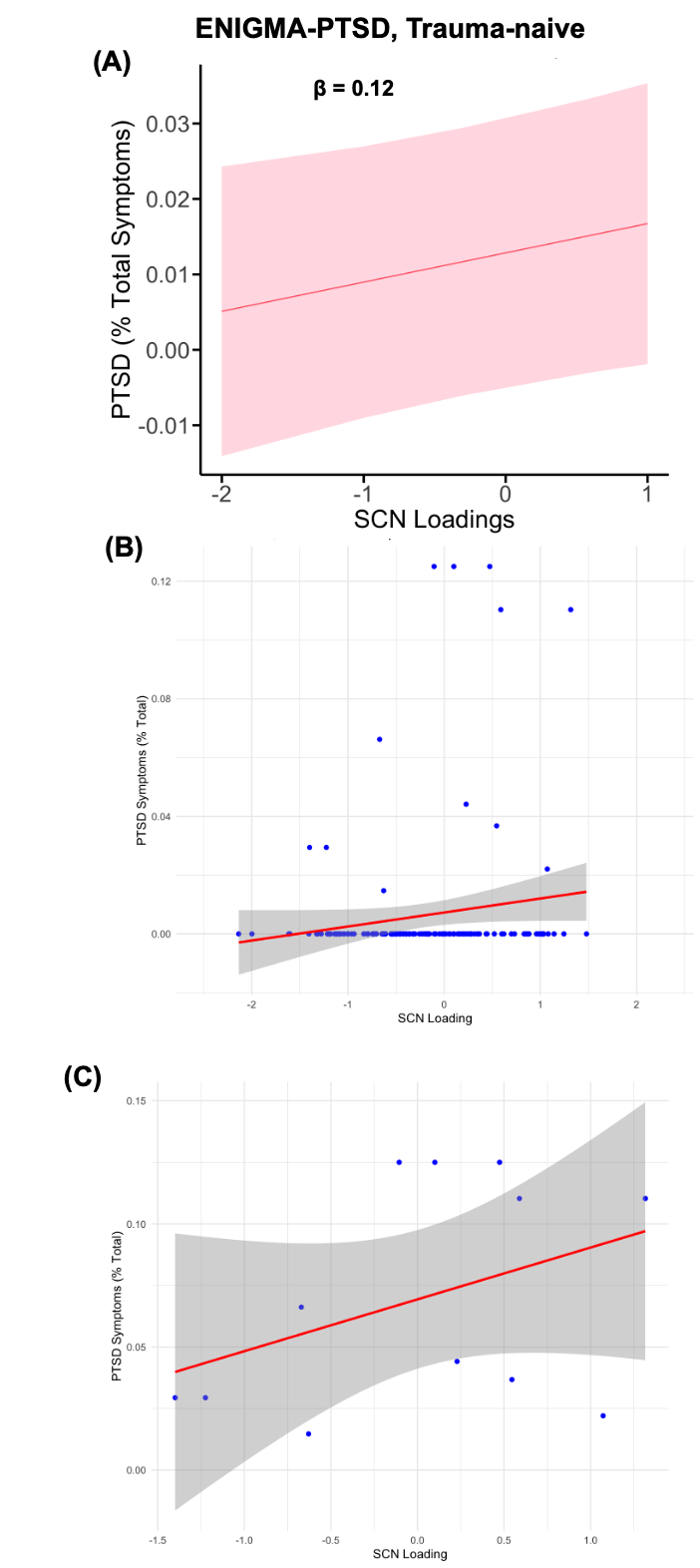

**Figure S3.** Violin plots show the distribution of structural covariance network (SCN) loadings for the trauma-naïve (i.e., HC), trauma-exposed with PTSD (i.e., PTSD), and trauma-exposed without PTSD (i.e., TEHC) within the ENIGMA-PTSD dataset.

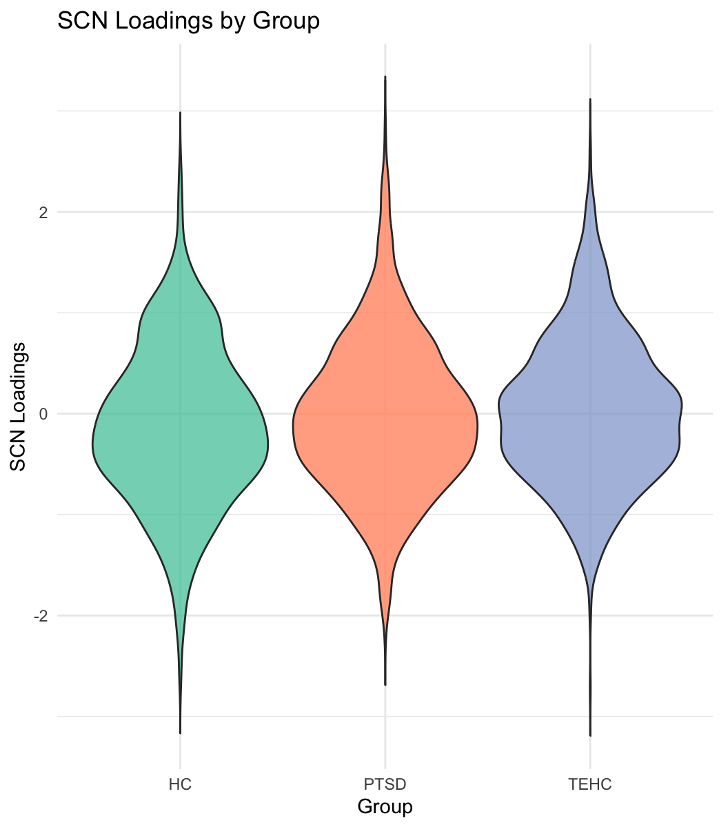

**Figure S4.** Early visual structural covariance network (SCN) loadings and ASR Total Scores in the HCP-YA dataset. The graph in (A) represents the partial plots from linear mixed effects models. Solid lines represent the unique association from the model and the shaded bars represent the confidence intervals. The graph in (B) represents the scatter plot for the independent variables. Solid lines represent the linear line of best fit, the shaded bars represent the confidence intervals, and blue dots represent the individual data points.

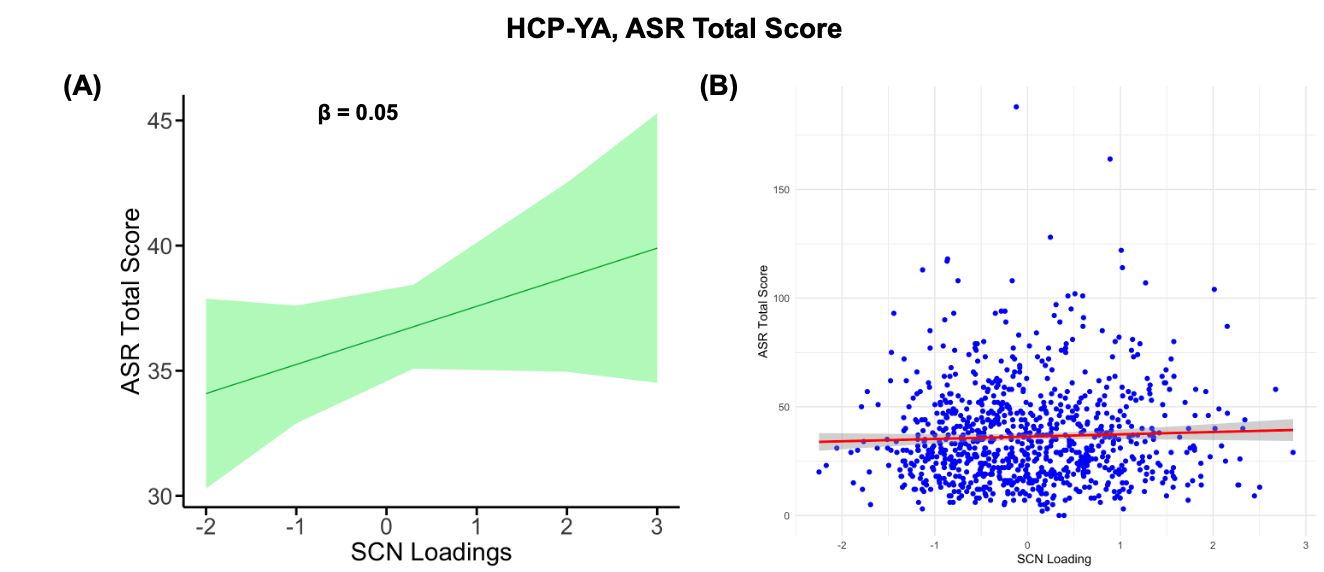
